# Adjunctive Corticosteroids for COVID-19: A Retrospective Cohort Study

**DOI:** 10.1101/2020.07.18.20157008

**Authors:** Ooi Say Tat, Purnima Parthasarathy, Lin Yi, Valliammai D/O Nallakaruppan, Shereen Ng Jia Huey, Tan Teck Choon, Low Serena, Tang Terence

## Abstract

**Background:** Coronavirus disease 2019 (COVID-19) is associated with severe pneumonia, respiratory failure and death. We aim to evaluate the efficacy of adjunctive corticosteroids in the management of COVID-19.

**Methods:** This is a retrospective cohort study of hospitalized adults (≥18 years) who were diagnosed with COVID-19 and were given treatment. Treatment included hydroxycholoroquine and lopinavir-ritonavir. Corticosteroids were included as adjunctive therapy in mid-April, 2020. We compared composite outcomes of clinical progression and invasive mechanical ventilation (MV) or death between group that received treatment only (Group A) versus group that received adjunctive corticosteroids (Group B). Entropy balancing was used to generate stabilized weight for covariates between treatment groups. Unweighted Kaplan-Meir curves, weighted and adjusted Cox regression analysis were used to estimate effect of adjunctive corticosteroids on composite outcomes. Subgroup analysis was performed on those with pneumonia.

**Results:** Of 1046 patients with COVID-19, 57 received treatment alone (Group A) and 35 received adjunctive corticosteroids in addition to treatment (Group B). Median day of illness at treatment initiation was 5 day. There were 44 patients with pneumonia; 68.9% of them were not requiring supplemental oxygen at treatment initiation. Overall, 17 (18.5%) of 92 patients had clinical progression including 13 (22.8%) of 57 patients in Group A versus 4 (11.4%) of 35 patients in Group B (p=0.172). Unweighted Kaplan-Meier estimates showed no significant difference in the proportion of patients who had clinical progression or invasive MV or death between the 2 treatment groups. However in those with pneumonia, there were lower proportions of patients in Group B with clinical progression (11.1%, 95% CI 0.0 - 22.2 versus 58.8%, 95% CI 27.3 - 76.7, log rank p<0.001); and invasive MV or death (11.3%, 95% CI 0.0 - 22.5 versus 41.2%, 95% CI 12.4. - 60.5, log rank p=0.016). In weighted and adjusted cox regression analysis, patients in Group B were less likely to have clinical progression, (adjusted HR [aHR] 0.08, 95% CI 0.01-0.99, p=0.049) but there was no statistical significant difference in risk of requiring invasive MV or death (aHR 0.22, 95%CI 0.02 - 2.54, p=0.22). In subgroup with pneumonia, patients in Group B were significantly at lower risk of clinical progression (aHR 0.15, 95% CI 0.06 - 0.39, p<0.001) and requiring invasive MV compared to Group A (aHR 0.30, 0.10-0.87, p=0.029).

**Conclusions:** Use of adjunctive corticosteroids is associated with lower risk of clinical progression and invasive MV or death, especially in those with pneumonia. Concurrent use of antivirals and corticosteroids should be considered in the management of COVID-19 related pneumonia.

## INTRODUCTION

Coronavirus disease 2019 (COVID-19) emerged in Wuhan city, China in December 2019 and has since evolved to a worldwide pandemic. Its clinical features range from asymptomatic to severe pneumonia requiring invasive mechanical ventilation and death.^1,2,3^

Severe disease is typically marked by rapid clinical deterioration with features of hyperinflammation.^4,5^

Considering its pathophysiology, use of immunomodulatory agent and corticosteroids have been proposed.^6^ Recent studies on use of tocilizumab and dexamethasone have shown promising results in reducing mortality in severe disease.^7,8^ However, treatment approach for early infection to prevent clinical progression to severe disease remains unclear.

At our center, decision to treat and treatment selection have evolved over time. It included interferon beta, lopinavir/ritonavir, hydroxycholoroquine (HCQ).

Prednisolone and tocilizumab were included as part of the treatment for COVID-19 in mid-April 2020 when there was reported hyperinflammation related to COVID-19 and use of anti-inflammatory agent might improve clinical outcome. ^9,10,11,12^ We aim to evaluate the effect of adjunctive corticosteroids in the management of COVID-19.

## METHODS

### Study design and participants

This is a retrospective, observational cohort study of patients with COVID-19 admitted to our hospital from February 9, 2020 to May 31, 2020.Data was retrieved from electronic medical records and included basic demographics, comorbidities, laboratory investigations and medications administered.

We included adults (≥18 years) with COVID-19, confirmed by PCR on nasopharyngeal swab who were admitted to our center and received treatment for COVID-19. Those who were on supportive care only, on invasive mechanical ventilation prior to treatment or those admissions not related to respiratory illness were excluded.

This study was approved by Domain Specific Review Board.

### Treatment

Decision to start treatment was at discretion of the treating physicians in consultation with an Infectious Diseases physician. Treatment regimen included lopinavir/ritonavir 400/100 mg twice a day, hydroxychloroquine 400mg twice on day 1, followed by 200mg three times a day on days 2-5 and prednisolone. Dose of prednisolone varied from 30mg once a day to 40mg twice a day at the physician’s discretion depending on severity of illness and patient’s body weight. Corticosteroids use of 3 days or more for any medical indications was considered as adjunctive corticosteroids.

Patients were classified into 2 groups, group that received usual treatment without corticosteroids (Group A) and group that received adjunctive corticosteroids in addition to usual treatment (Group B). Clinical conditions of the patients with COVID-19 included at initiation of treatment were categorized into 3 stages: COVID-19 without pneumonia (Stage 1), COVID-19 related pneumonia not requiring supplemental oxygen (Stage 2) and COVID-19 related pneumonia requiring supplemental oxygen (Stage 3). Chest radiograph was the primary modality used in the assessment for pneumonia in our cohort.

### Outcomes

Primary outcome was a composite of clinical progression or death. Clinical progression is defined as progression to requirement of supplemental oxygen in Stage 1 and 2 or progression to invasive mechanical ventilation in any stage whichever is earlier. Secondary outcome was a composite of invasive mechanical ventilation or death.

Subgroup analysis was performed on those with pneumonia (Stage 2 & 3)

### Statistical analysis

Categorical variables are shown as numbers and percentages while continuous variables as median with interquartile range (IQR). Comparison of continuous and categorical data between groups was done using Mann-Whitney U test and χ2 test or Fisher’s exact test as appropriate.

Magnitude of changes in C-reactive protein (CRP) level following initiation of treatment were calculated using available CRP values compared to baseline CRP (delta CRP). Median values of delta CRP and daily maximum temperature following initiation of treatment up to 14 days were plotted by treatment group for comparison.

Data was preprocessed with entropy balancing to achieve covariate balance between the two treatment groups to reduce model dependence for the subsequent estimation of treatment effects.^13^ Weight derived from maximum entropy reweighting scheme was used to generate stabilized weight for each subject in the treatment groups. Balanced covariate is defined by standardized mean difference of less than 0.1 and variance ratio between 0.5 - 2.0. Procedure of covariate balance was performed separately for all eligible subjects and in subgroup with pneumonia. Baseline covariates used to assess balanced weighting between treatment groups were age, gender, body mass index (BMI), comorbidity, clinical stage, antibiotic use, CRP, procalcitonin, lymphocyte count, neutrophils-lymphocyte ratio and creatinine.

Figure 2 displays the standardized mean difference of overall cohort subjects and subgroup with pneumonia between the two treatment groups. Imbalanced covariates between the two treatment groups were age, procalcitonin and creatinine. In subgroup with pneumonia, age, procalcitonin, creatinine and neutrophil-lymphocyte ratio were imbalanced (Table S1).

**Figure 1:**
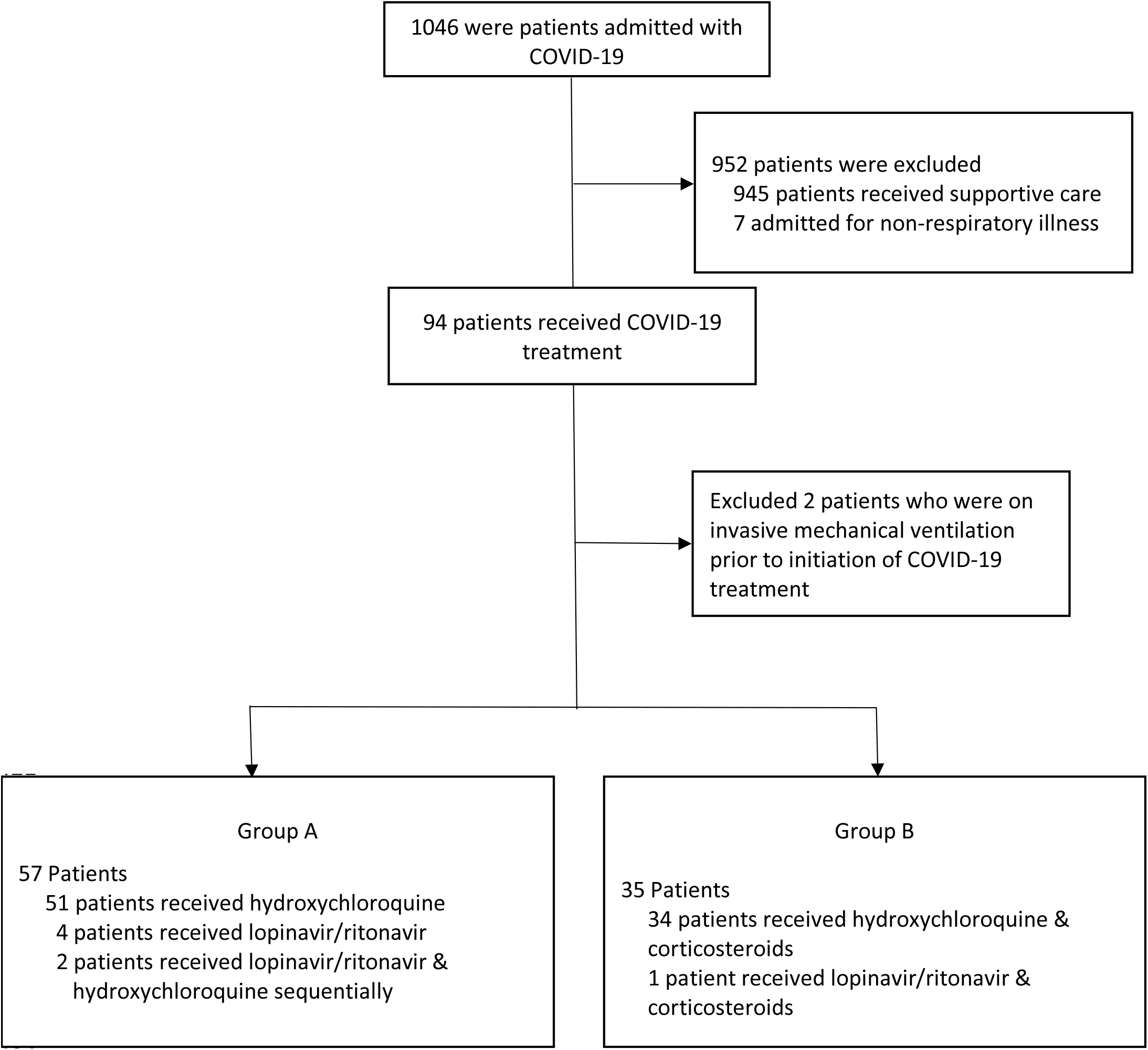
Overview of patients included in the study cohort.

**Figure 2.**
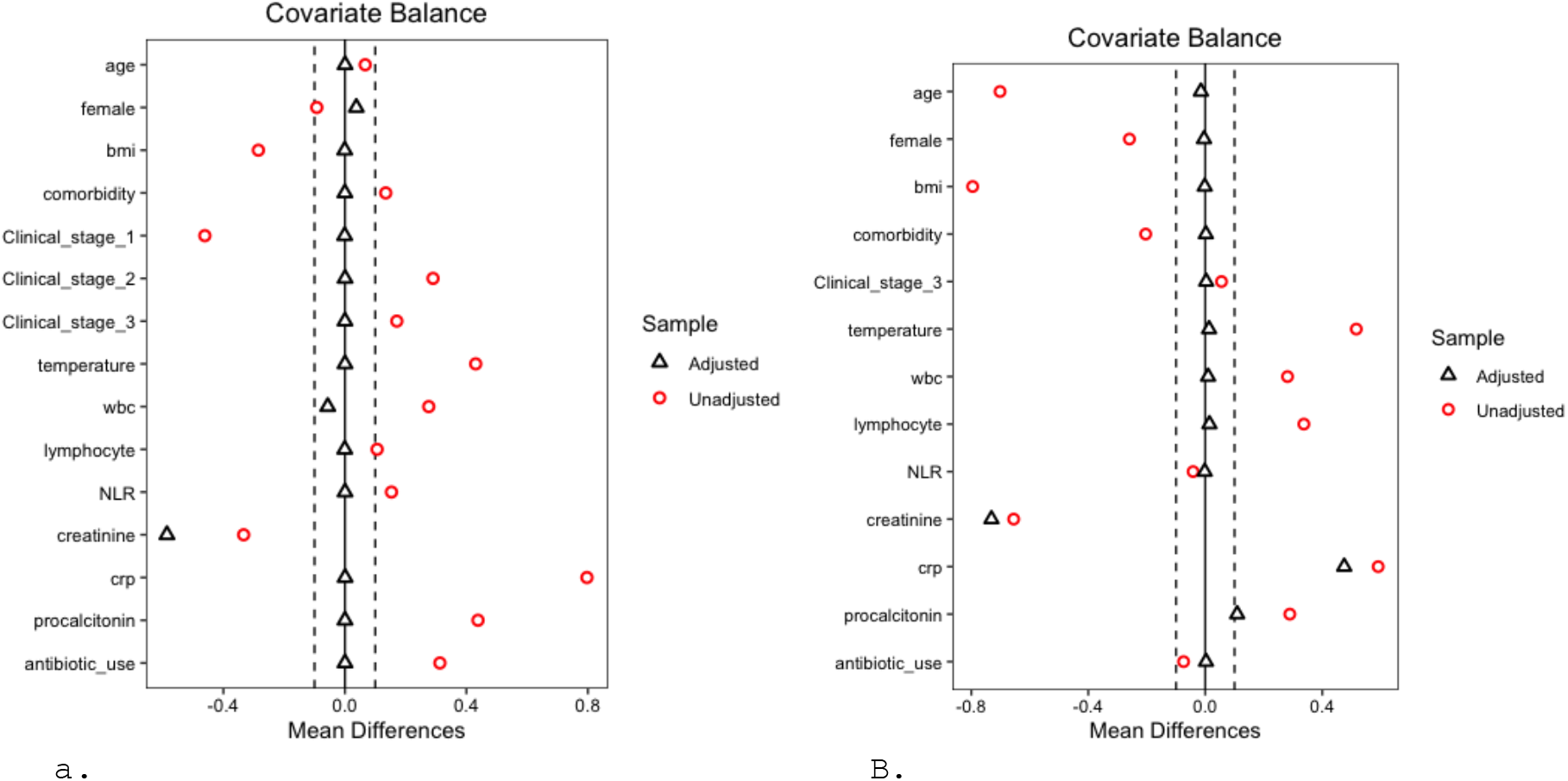
Adjusted and unadjusted covariate balance in overall cohort(a) and in subgroup with pneumonia(b) between two treatment groups. Temperature is the highest recorded body temperature on the day of treatment initiation. bmi, body mass index; wbc, white blood cells; NLR, neutrophil-lymphocyte ratio; CRP, C-reactive protein.

Missing data of covariates were mainly limited to CRP and procalcitonin (1.1%). Multiple imputation was applied for missing data.

Kaplan Meier curves and cox regression analysis were used to compare time to composite outcomes between 2 treatment groups. Patients without primary end-point event were censored on date of discharge. Proportions of patients with composite outcomes between 2 treatment groups were compared using unweighted Kaplan-Meier estimates. We used weighted Cox proportional-hazards regression models with adjustment for imbalanced covariates to estimate effect of adjunctive corticosteroids on composite outcomes. The effect estimate is shown using hazard ratio (HR) with 95% confidence interval (95% CI).

We considered two-sided p value test of less than 0.05 as statistically significant. Comparison of baseline characteristics by treatment group, patterns of fever and CRP changes following treatment was performed using STATA version 15 (STATA Corp., Texas, USA). Multiple imputation, entropy balancing with covariate balance assessment and Cox regression analyses were performed using R Statistical Software (Foundation for Statistical Computing, Vienna, Austria) and related R packages (MICE, WeighIt, ebal and Cobalt).^14,15^

## RESULTS

Between February 9, 2020 and May 31, 2020, there were 1046 patients with covid-19 admitted to our hospital. Of these, 94 (9.0%) patients received treatment. Two patients were excluded as treatment was started following invasive mechanical ventilation. In total, 57 patients received treatment without corticosteroids (Group A) and 35 patients received adjunctive prednisolone to treatment (Group B). (Figure 1)

Median number of days of illness at the start of treatment was 4 days and 5 days in Group A and B respectively. Duration of HCQ used was 7 days (IQR 5-7) in Group A and 5 days (IQR 5-7) in Group B.

Prednisolone use was 5 days (IQR 3-7).

There were 44 patients in the cohort who had pneumonia. Of these, 68.9% of them were not requiring supplemental oxygen at treatment initiation. Between the 2 groups, there were higher proportions of patients in Group B who had pneumonia and requiring supplemental oxygen, use of antibiotics and higher CRP level at initiation of treatment (Table 1). Eight subjects received tocilizumab in our cohort. All were given post mechanical ventilation except 2 cases (one in each treatment group).

**Table 1:** Baseline characteristics of overall cohort by treatment group.

|  | Group A<br>N=57 (%) | Group B<br>N=35 (%) | P-value |
| --- | --- | --- | --- |
| Age (year) | 50 (36-58) | 48 (41-57) | 0.94 |
| Female | 12(21%) | 4(11%) | 0.24 |
| Ethnicity |  |  | 0.10 |
| Chinese | 12 (21%) | 14 (40%) |  |
| Malay | 10 (18%) | 1(3%) |  |
| Indian | 17(30%) | 8(23%) |  |
| Others | 18 (31%) | 12 (34%) |  |
| Body mass index (kg/m<sup>2</sup>) | 25.5 (23.4-28.4) | 25.6 (21.7-26.7) | 0.40 |
| Comorbidity present | 26(46%) | 21(60%) | 0.18 |
| Diabetes mellitus | 14 (25%) | 14 (40%) | 0.12 |
| Hypertension | 16 (28%) | 6 (17%) | 0.23 |
| Hyperlipidaemia | 7 (12%) | 3 (9%) | 0.58 |
| Renal diseases | 10 (18%) | 5 (14%) | 0.68 |
| Cardiac diseases | 3 (5%) | 2 (6%) | 0.93 |
| Neurological diseases | 0 (0%) | 1 (3%) | 0.20 |
| Rheumatologic diseases | 2 (4%) | 1 (3%) | 0.86 |
| Respiratory diseases | 1 (2%) | 1(3%) | 0.72 |
| Day of Illness* | 4 (2, 6) | 5 (4, 9) | 0.064 |
| Clinical Stage upon treatment |  |  | <0.001 |
| Stage 1 | 40 (70%) | 8 (23%) |  |
| Stage 2 | 13 (23%) | 18 (51%) |  |
| Stage 3 | 4 (7%) | 9 (26%) |  |
| Max Temperature (°C)** | 38.5 (38.1-39.1) | 38.8 (38.4-39.3) | 0.06 |
| Use of antibiotics before treatment | 14 (25%) | 20 (57%) | 0.002 |
| Use of inotropes | 2(4%) | 0 (0%) | 0.26 |
| Use of tocilizumab | 3(5%) | 5 (14%) | 0.14 |
| C-reactive protein (mg/L) | 12.7 (6.2-33.9) | 60.7 (37.9-109.3) | <0.001 |
| Procalcitonin (ng/mL) | 0.10 (0.06- 0.15) | 0.13 (0.08-0.27) | 0.039 |
| White blood cells (x10<sup>9</sup>/L) | 5.5 (4.6- 6.9) | 6.3 (4.1, 8.9) | 0.27 |
| Lymphocytes (x10<sup>9</sup>/L) | 1.2 (0.9-1.6) | 1.2 (0.9-1.6) | 0.95 |
| Neutrophils (x10<sup>9</sup>/L) | 3.7 (2.6- 4.8) | 4.7(2.9-6.4) | 0.05 |
| Monocytes (x10<sup>9</sup>/L) | 0.51 (0.36-0.83) | 0.37 (0.27-0.51) | 0.004 |
| Creatinine (umol/L) | 84 (73-94) | 74 (67-87) | 0.06 |
| Bicarbonate (mmol/L) | 24 (23-26) | 24 (23-25) | 0.54 |
| Aspartate aminotransferase (U/L) | 30 (21-37) | 33 (23-44) | 0.24 |
| Alanine aminotransferase (U/L) | 28 (22-37) | 32 (20-41) | 0.63 |

Data are median (IQR) or n(%) unless otherwise stated. P values were calculated using Mann-Whitney U test and $\chi^2$ test or Fisher's exact test as appropriate.
\*Day of illness represent the number of day of illness by symptoms on the day of treatment initiation.
\*\*Max temperature is the highest recorded body temperature on the day of treatment initiation.

Following treatment initiation, body temperature of patients trended down in similar pattern between the 2 treatment groups. CRP in Group B trended down. However, in Group A, it trended up at around 5-8 days following treatment before declining (Figure 3).

**Figure 3:**
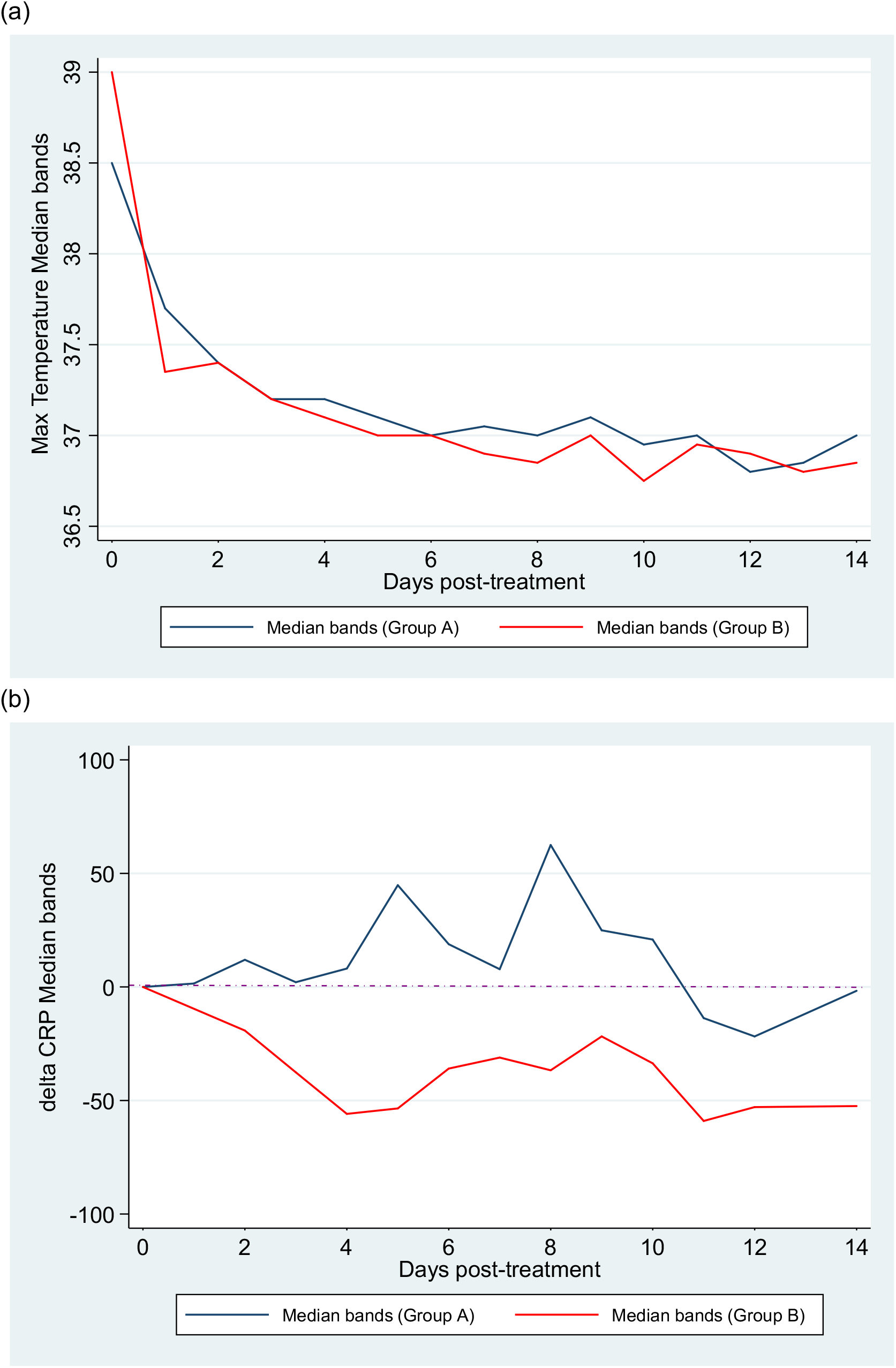
Trends of fever and changes of C-reactive protein level by treatment groups. (a) Note: delta C-reactive protein (CRP) is the changes in CRP level following initiation of treatment were calculated compared to baseline CRP level at initiation of treatment

Overall, 17 (18.5%) of 92 patients had clinical progression including 13 (22.8%) of 57 patients in Group A versus 4 (11.4%) of 35 patients in Group B (p=0.172) (Table S2 in supplementary material).

There were 11 patients required invasive mechanical ventilation; 7 (12.2%) patients in Group A and 4 (11.4%) patients in Group B. One death occurred in Group A following invasive mechanical ventilation and none in Group B.

Unweighted Kaplan-Meier estimates showed no significant difference in the proportion of patients who had clinical progression or invasive mechanical ventilation between the 2 treatment groups (Figure 4). At day 12 after treatment initiation, the proportion of patients with clinical progression was 23.3% (95% CI 11.3 - 33.7) in Group A versus 11.4% (95% CI 0.2 - 21.4) in Group B (log rank p=0.16); 12.3% (95% CI 3.3. - 20.4) of patients in Group A required invasive mechanical ventilation versus 12.2% (95% CI 0.2 - 22.7) of patients in Group B (log rank p=0.85). However, in subgroup analysis of those with pneumonia, there was beneficial effect of adjunctive prednisolone compared to treatment alone. At day 12 after treatment initiation, the proportion of patients with clinical progression was 58.8% (95% CI 27.3 - 76.7) in Group A versus 11.1% (95% CI 0.0 - 22.2) in Group B (log rank p<0.001); 41.2% (95% CI 12.4. - 60.5) of patients in Group A required invasive mechanical ventilation versus 11.3% (95% CI 0.0 - 22.5) of patients in Group B (log rank p=0.016).

**Figure 4.**
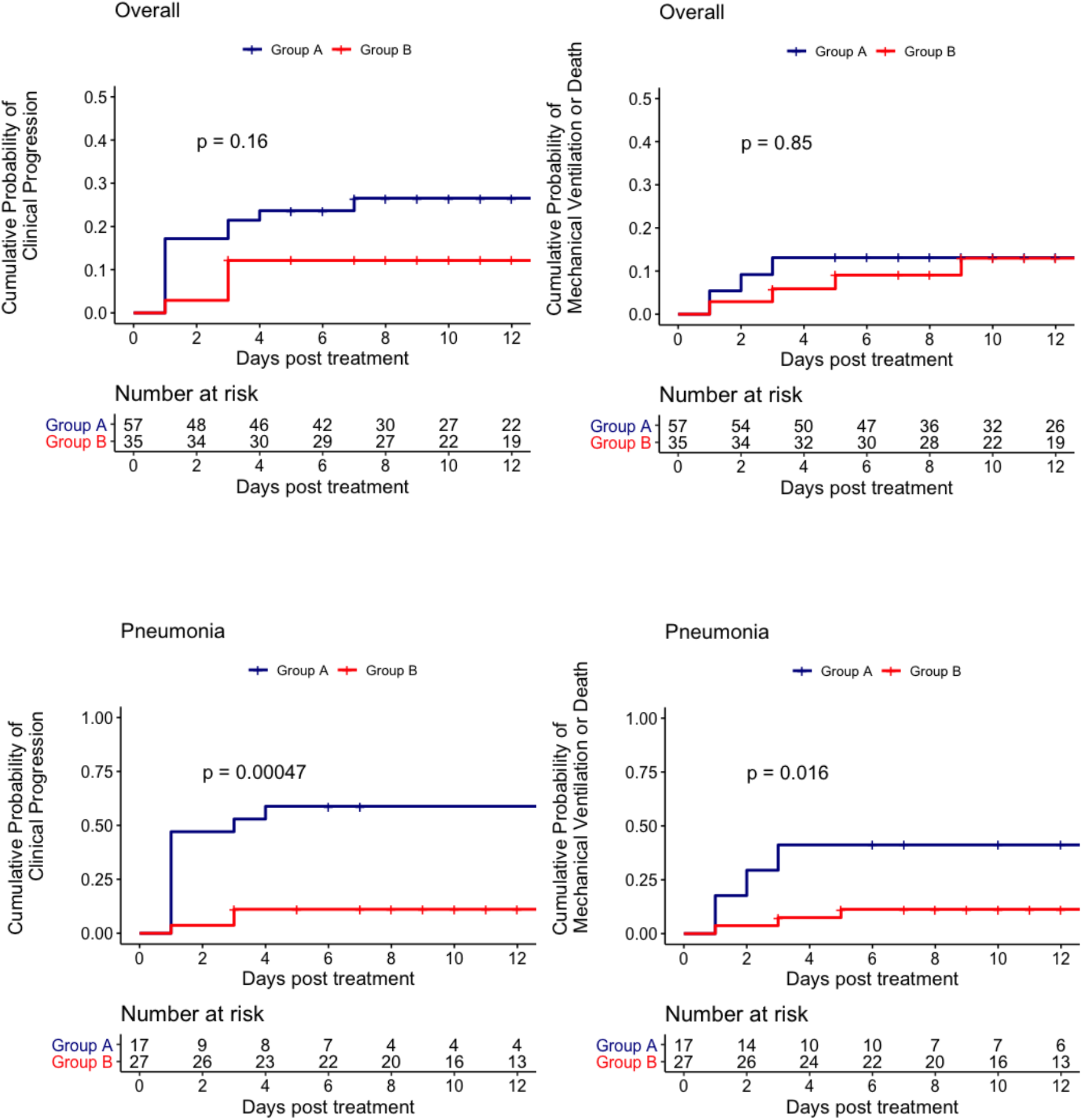
Kaplan-Meier estimates of the cumulative probability of clinical progression and mechanical ventilation or death by treatment group.

In unadjusted cox regression, there was no significant difference in risk of clinical progression between the two treatment groups (Table 3). In weighted and adjusted cox regression analysis, patients in Group B were less likely to have composite outcome of clinical progression compared to Group A, (adjusted HR [aHR] 0.08, 95% CI 0.01-0.99, p=0.049) but there was no statistical significant difference in risk of requiring invasive mechanical ventilation or death (aHR 0.22, 95%CI 0.02 - 2.54, p=0.22). In subgroup analysis of those with pneumonia, patients in Group B were significantly at lower risk of clinical progression (aHR 0.15, 95% CI 0.06 - 0.39, p<0.001) and requiring invasive mechanical ventilation compared to Group A (aHR 0.30, 0.10-0.87, p=0.029).

**Table 3.** Unadjusted and adjusted relative hazards of clinical progression and Invasive mechanical ventilation.

|  | N | Unadjusted analysis<br>HR (95% CI) | p value | Adjusted analysis<br>HR (95% CI) | p value |
| --- | --- | --- | --- | --- | --- |
| **Clinical Progression** |  |  |  |  |  |
| Overall | 92 | 0.46(0.15 – 1.40) | 0.169 | 0.08 (0.01 – 0.99) | 0.049 |
| Pneumonia | 44 | 0.13(0.04 – 0.48) | 0.002 | 0.15 (0.06 – 0.39) | <0.001 |
| **Invasive Mechanical Ventilation** |  |  |  |  |  |
| Overall | 92 | 0.88 (0.26 – 3.02) | 0.844 | 0.22 (0.02 – 2.54) | 0.22 |
| Pneumonia | 44 | 0.22 (0.06 – 0.85) | 0.028 | 0.30 (0.10 – 0.87) | 0.029 |

N, number; HR, hazard ratio; CI, confidence interval.
For adjusted HR in overall group, age, C-reactive protein, procalcitonin were adjusted using weighted Cox regression analysis.
For adjusted HR in pneumonia, age, C-reactive protein, procalcitonin and neutrophil-lymphocyte ratio were adjusted using weighted Cox regression analysis.

## DISCUSSION

Severe acute respiratory syndrome coronavirus 2 (SARS-Cov-2) infection is characterized by high viral replication during the initial phase of illness and a robust hyper immune response leading to morbidity and mortality. ^9,10,16^ Our study provides important insights into the management of early COVID-19-related pneumonia, particularly highlighting the need to address both the viral replication and the inflammatory response in the early stages of infection.

Our cohort of patients either received antiviral alone (mostly HCQ) or were treated with a combination of HCQ and corticosteroids (Prednisolone) during the early phase of illness. The use of adjunctive prednisolone along with HCQ was associated with a significantly lowered risk of clinical disease progression, especially in those with pneumonia. 68.9% of patients with pneumonia in our cohort were not on supplemental oxygen at initiation of treatment. Early use of prednisolone in our patients with COVID-19 pneumonia and not on supplemental oxygen, had a 70% lower risk of progressing to invasive mechanical ventilation or death compared to non-steroid group (HR 0.30, 95% CI 0.10 - 0.87). The duration of prednisolone use in our cohort was short, with a median of 5 days. The dose of prednisolone varied between 0.5-1.0 mg/kg/day.

Emerging data have shown benefits of anti-inflammatory agents in the management of severe COVID-19 pneumonia, especially with the use of tocilizumab and corticosteroids.^7,8^ The RECOVERY trial involving the use of dexamethasone in hospitalized patients with COVID-19 in UK showed a 20%-40% reduction in mortality in those with pneumonia requiring oxygen and invasive mechanical ventilation. However, there was a trend towards increased mortality when corticosteroids were used in patients not receiving respiratory support at randomization.^8^ This seems to correspond to use of corticosteroids in early pneumonia. Earlier observations of adverse outcomes with corticosteroids use in SARS and Middle East respiratory syndrome (MERS-CoV) infections may explain the undesired consequences of corticosteroids in early phase of COVID-19 pneumonia at which virus replication is active.^17^

HCQ has been shown to effectively inhibit SARS-CoV-2 infection in vitro.^18^ Despite its in-vitro activity, clinical efficacy of HCQ has largely been reported as ineffective. The RECOVERY trial reported an increased risk of invasive mechanical ventilation or death with use of HCQ compared to supportive care.^19^ We found that the group on treatment with antivirals without adjunctive corticosteroids in our cohort had a more protracted course with a higher inflammatory response. Their CRP trended to higher level at day 5-8 post treatment despite a similar fever response in both groups. Perhaps this is a paradoxical inflammatory reaction due to treatment. We might draw simile to post-treatment inflammatory response in *pneumocystis jiroveci* pneumonia, where early adjunctive corticosteroids could decrease risk of respiratory failure.^20^

The use of prednisolone during early pneumonia in our study had a significantly better outcome which may be due to the concurrent administration of antiviral agents such as HCQ or lopinavir/ritonavir. A similar observation could be from a retrospective study that showed significant lower risk of invasive mechanical ventilation or death with the use of tocilizumab to standard care that included HCQ and lopinavir/ritonavir in the first week of COVID-19 pneumonia.^7^

The prominent protective effect of adjunctive prednisolone observed in our cohort with early pneumonia and the varying effect of dexamethasone on different days of illness of COVID-19 in the RECOVERY trial suggest the need for different treatment strategies at different clinical stages of COVID-19 infection. Our results support the hypothesis that control of both viral replication and inflammatory response is important in treatment of early COVID-19 pneumonia.^5^

Our study has several limitations. Without a placebo arm, we are unable to conclude if the protective effect observed in our study was directly due to corticosteroids alone or in combination with a treatment agent, mostly HCQ. Non-randomized treatment selection and small sample size could have biased estimates and limit our subgroup analysis. The estimates in weighted regression are dependent on balancing of covariates and confounders.

Despite non-parametric method of covariate balancing, there were some imbalances that required further covariate adjustment in regression models. Our estimates could subject to possible model misspecification and are based on assumption that all confounders are controlled. In a conservative view, estimates in unadjusted cox regression did not show an increased risk of composite outcomes despite seemingly more severe disease in group that received adjunctive corticosteroids. Having said that, risk of secondary infection and other adverse outcomes associated with corticosteroids were not evaluated.

Our analysis could not account for change in clinical practice and experience gained in managing COVID-19 over time that might affect better outcome in the later stage. However, it is unlikely to account for lower risk of clinical progression of requiring oxygen in those with pneumonia on adjunctive corticosteroids.

Lastly, younger population in our cohort and single center design may limit generalizability of these results to other populations.

In summary, our study shows that the use of adjunctive corticosteroids was associated with lower risk of clinical progression and invasive MV or death, especially in those with pneumonia. Patients in early phase of COVID-19 pneumonia may benefit from adjunctive corticosteroid therapy. Further prospective studies are required to evaluate the clinical efficacy of a combination treatment with antivirals and anti-inflammatory agents in early COVID 19 pneumonia.

## Data Availability

NA

## Supplementary Material

**Table S1a.** Covariate balance of the overall cohort between two treatment groups

| Covariate | Type | Unadjusted difference* | Unadjusted Variance Ratio | Adjusted difference* | Adjusted Variance Ratio |
| --- | --- | --- | --- | --- | --- |
| Age | Continuous | 0.0668 | 0.8349 | 0.0005 | 0.351 |
| Female | Binary | -0.0926 | NA | 0.0376 | NA |
| BMI | Continuous | -0.2848 | 0.6576 | -0.0005 | 0.9822 |
| Comorbidity | Binary | 0.1345 | NA | -0.0001 | NA |
| Stage_1 | Binary | -0.4611 | NA | -0.0007 | NA |
| Stage 2 | Binary | 0.2901 | NA | 0.0005 | NA |
| Stage 3 | Binary | 0.1709 | NA | 0.0002 | NA |
| Temperature** | Continuous | 0.4306 | 0.8152 | 0.0003 | 0.6452 |
| White cells | Continuous | 0.2761 | 1.7111 | -0.0565 | 1.8464 |
| Lymphocytes | Continuous | 0.1061 | 1.5234 | -0.0005 | 1.8224 |
| Neutrophils-lymphocyte ratio | Continuous | 0.1533 | 1.1089 | 0.0004 | 1.4704 |
| Creatinine | Continuous | -0.3333 | 0.8675 | -0.5859 | 0.6754 |
| C-reactive protein | Continuous | 0.7969 | 2.304 | 0.0007 | 0.5673 |
| Procalcitonin | Continuous | 0.4382 | 8.9343 | 0.0003 | 2.8179 |
| Antibiotic use | Binary | 0.3128 | NA | 0.0003 | NA |

**Table S1b.** Covariate balance of subgroup with pneumonia between two treatment groups

| Covariate | Type | Unadjusted difference* | Unadjusted Variance Ratio | Adjusted difference* | Adjusted Variance Ratio |
| --- | --- | --- | --- | --- | --- |
| Age | Continuous | -0.7017 | 1.6742 | -0.0149 | 2.0342 |
| Female | Binary | -0.2593 | NA | -0.0043 | NA |
| BMI | Continuous | -0.7953 | 0.6944 | -0.0018 | 0.9756 |
| Comorbidity | Binary | -0.2037 | NA | 0.002 | NA |
| Stage_1 | Binary | -0.0556 | NA | -0.0031 | NA |
| Stage 2 | Binary | 0.0926 | NA | -0.0012 | NA |
| Stage 3 | Binary | -0.037 | NA | 0.0043 | NA |
| Temperature** | Continuous | 0.5164 | 0.866 | 0.013 | 0.719 |
| WBC | Continuous | 0.2812 | 1.1032 | 0.0097 | 0.574 |
| Lymphocytes | Continuous | 0.3371 | 1.8044 | 0.0143 | 1.8283 |
| Neutrophils-lymphocyte ratio | Continuous | -0.0416 | 0.3842 | -0.0019 | 0.2974 |
| Creatinine | Continuous | -0.655 | 0.3425 | -0.7311 | 0.2618 |
| CRP | Continuous | 0.5912 | 2.5667 | 0.475 | 1.6594 |
| Procalcitonin | Continuous | 0.2891 | 5.1193 | 0.1091 | 4.1444 |
| Antibiotic use | Binary | -0.0741 | NA | 0.0025 | NA |

**Table S2.** Composite outcomes by clinical stages and treatment groups

| Composite of Clinical progression | Stage 1 |  | Stage 2 |  | Stage 3 |  |
| --- | --- | --- | --- | --- | --- | --- |
|  | Group A (n=40) | Group B (n=8) | Group A (n=13) | Group B (n=18) | Group A (n=4) | Group B (n=9) |
| Supplemental oxygen only | 3 (7.5%) | 1 (12.5%) | 7 (53.8%) | 1 (5.6%) | NA | NA |
| Invasive mechanical ventilation | 0 (0%) | 1 (12.5%) | 4 (30.8%) | 1 (5.6%) | 3 (75%) | 2 (22.2%) |

Stage 1, no pneumonia; Stage 2, pneumonia with no supplemental oxygen; Stage 3, pneumonia on supplemental oxygen

## References

1. Rothe C, Schunk M, Sothmann P, et al. Transmission of 2019-nCoV infection from an asymptomatic contact in Germany. N Engl J Med 2020;382:970–971.

2. Guan WJ, Ni ZY, Hu Y, et al. Clinical Characteristics of Coronavirus Disease 2019 in China. N Engl J Med 2020;382:1708–20.

3. Wang D, Hu B, Hu C, et al. Clinical Characteristics of 138 Hospitalized Patients With 2019 Novel Coronavirus-Infected Pneumonia in Wuhan, China. JAMA 2020.

4. Pedersen SF, Ho YC. SARS-CoV-2: a storm is raging. J Clin Invest. 2020;130(5):2202–2205.

5. Siddiqi HK, Mehra MR. COVID-19 illness in native and immunosuppressed states: A clinical-therapeutic staging proposal. J Heart Lung Transplant. 2020;39(5):405–407.

6. Mehta P, McAuley DF, Brown M, et al. COVID-19: consider cytokine storm syndromes and immunosuppression. Lancet. 2020;395(10229):1033–1034.

7. Guaraldi G, Meschiari M, Cozzi-Lepri A, et al. Tocilizumab in patients with severe COVID-19: a retrospective cohort study. Lancet Rheumatol 2020; published online June 24. https://doi.org/10.1016/S2665-9913(20)30173-9

8. The RECOVERY Collaborative Group. Dexamethasone in hospitalized patients with Covid-19 — preliminary report. N Engl J Med. 2020. DOI: 10.1056/NEJMoa2021436.

9. Huang C, Wang Y, Li X, et al. Clinical features of patients infected with 2019 novel coronavirus in Wuhan, China. Lancet 2020; 395: 497–506.

10. Zhou F, Yu T, Du R, et al. Clinical course and risk factors for mortality of adult inpatients with COVID-19 in Wuhan, China: a retrospective cohort study. Lancet. 2020;395(10229):1054–1062.

11. Wu C, Chen X, Cai Y, et al. Risk factors associated with acute respiratory distress syndrome and death in patients with coronavirus disease 2019 pneumonia in Wuhan, China. JAMA Intern Med 2020,180(7):–11.

12. Luo P, Liu Y, Qiu L, Liu X, Liu D, Li J. Tocilizumab treatment in COVID-19: A single center experience. J Med Virol. 2020;92(7):814–818.

13. Hainmueller J. Entropy Balancing for Causal Effects: A Multivariate Reweighting Method to Produce Balanced Samples in Observational Studies. Political Analysis. 2012;20(1):25–46

14. van Buuren S, Groothuis-Oudshoorn K (2011). mice: Multivariate Imputation by Chained Equations in R. Journal of Statistical Software, 45(3),1–67.

15. Greifer, N. cobalt: Covariate Balance Tables and Plots. R package version 4.2.2 (2020)

16. To KK, Tsang OT, Leung WS, et al. Temporal profiles of viral load in posterior oropharyngeal saliva samples and serum antibody responses during infection by SARS-CoV-2: an observational cohort study. Lancet Infect Dis. 2020;20(5):565–574.

17. Russell CD, Millar JE, Baillie, JK. Clinical evidence does not support corticosteroid treatment for 2019-nCoV lung injury. Lancet. 2020;395(10223):473–475.

18. Liu, J., Cao, R., Xu, M. et al. Hydroxychloroquine, a less toxic derivative of chloroquine, is effective in inhibiting SARS-CoV-2 infection in vitro. Cell Discov. 2020;6:16.

19. Horby P, Mafham M, Linsell L, et al. Effect of Hydroxychloroquine in Hospitalized Patients with COVID-19: Preliminary results from a multi-centre, randomized, controlled trial. medRxiv 2020. Available at: https://www.medrxiv.org/content/10.1101/2020.07.15.20151852v1

20. Gagnon S, Boota AM, Fischl MA, et al. Corticosteroids as adjunctive therapy for severe Pneumocystis carinii pneumonia in the acquired immunodeficiency syndrome: a double-blind, placebo-controlled trial. N Eng J Med 1990; 323:1444–1450

